# Pooled RNA sample reverse transcriptase real time PCR assay for SARS CoV-2 infection: a reliable, faster and economical method

**DOI:** 10.1101/2020.04.25.20079095

**Authors:** Ekta Gupta, Abhishek Padhi, Arvind Khodare, Reshu Aggarwal, Krithiga Ramachnadran, Vibha Mehta, Mousumi Kilikdar, Shantanu Dubey, Guresh Kumar, Shiv Kumar Sarin

**Author notes:** **Corresponding Author:** Prof. Shiv Kumar Sarin, Senior Professor, Hepatology, Institute of Liver and Biliary Sciences, Vasant Kunj, New Delhi, India. **Alternate corresponding author** Dr Ekta Gupta, Professor, Clinical virology, Institute of Liver and Biliary Sciences, Vasant Kunj, New Delhi, India.

## Abstract

**Background:** Corona virus disease 2019 (COVID-19) which initially started as a cluster of pneumonia cases in the Wuhan city of China has now become a full blown pandemic. Timely diagnosis of COVID-19 is the key in containing the pandemic and breaking the chain of transmission. In low and middle income countries availability of testing kits has become the major bottle neck in testing. Novel methods like pooling of samples are the need of the hour.

**Method:** Extracted RNA samples were randomly placed in pools of 8 on a 96 well plate. Both individual RNA (ID) and pooled RNA RT-qPCR for the screening E gene were done in the same plate and the positivity for the E gene was seen.

**Results:** The present study demonstrated that pool testing with 8 RNA samples can easily detect even up to a single positive sample with Ct value as high as 38. The present study also showed that the results of pool testing is not affected by number of positive samples in a pool.

**Conclusion:** Pooling of 8 RNA samples can reduce the time and expense by one eighth, and can help expand diagnostic capabilities, especially during constrained supply of reagents and PCR kits for the diagnosis of SARS-CoV-2 infection.

## INTRODUCTION

Corona virus disease 2019 (COVID-19) is a severe acute respiratory infection caused by the novel corona virus, severe acute respiratory syndrome coronavirus 2 (SARS-CoV-2) [1]. Initially started as cluster of cases from Wuhan, China [2] it has now spread to over 200 countries with 1,436,198 confirmed and 85, 522 deaths [3]. The laboratory diagnosis of SARS-CoV-2 is based primarily on nucleic acid amplification test (NAAT) like real time reverse transcriptase PCR (RT-qPCR). With an acute shortage of diagnostic kits the present testing strategies mainly focus on testing the symptomatic individuals. But detecting the carrier or asymptomatic individuals holds the key in containing the spread of the infection into the community. Earlier the infected person is identified, sooner the spread of the infection can be contained and the surveillance machinery can be activated for contact tracing and ultimately break in the chain of transmission of the virus. Most of the countries have imposed lockdown to contain the infection but afterwards when the cases are expected to take a vertical trajectory, scaling up the testing would be a major challenge [4]. Innovative methods to conserve kits and reagents and human resources needs to be explored to enhance the testing. Pooling the diagnostic tests has been applied in other infectious diseases and is especially attractive as it requires no additional training, equipment, or materials. In this method, perfected over the years [5] [6] [7], samples are mixed and tested at a single pool, and subsequent individual tests are performed, only if the pool tests positive.

We undertook this study to evaluate a novel protocol of pooling of RNA samples/elutes in performance of PCR for SARS CoV-2 virus. We were extremely careful that the limits of dilution, do not compromise the sensitivity and specificity of the results.

## METHODOLOGY

### Sample Collection

Combined nasopharyngeal and oropharyngeal swabs were collected by healthcare workers and transported in a 3 ml viral transport media(VTM) maintaining proper cold chain and sent to the virology laboratory of Institute of Liver and biliary sciences, New Delhi, India. A volume of 200 microlitre (µl) of the sample was further processed for viral nucleic acid extraction by Qiasymphony DSP Virus/ Pathogen mini kit (Qiagen GmbH, Germany) as per the manufacturers protocol in elutes of 60 µl each [8]. Each sample was subjected to the addition of 10 µl of spiked extraction control (EAC) at the time of extraction itself, to check the validity of the extraction procedure.

### PERFORMANCE OF RT-qPCR IN THE LABORATORY

The 5 µl of the extracted RNA elute/sample was subjected to RT-qPCR for the qualitative detection of SARS-CoV-2 RNA utilizing with AgPath-IDTM One-Step RT-PCR Reagents (Thermo Fisher Scientific) using a Applied biosystem (ABI) 7500 Real Time PCR system (Thermo Fisher Scientific) and LightMix® SarbecoV E-gene (TIB MOLBIOL). Reactions were heated to 55° C for 5 minutes for reverse transcription, denatured in 95° C for 5 minutes and then 45 cycles of amplification were carried out for 95°C for 5 seconds and 60°C for 15 seconds using FAM parameter for E gene. This assay targets the detection of E gene for SARS as well as nCoV-2. The primer details are given in Table 1. All samples that were screened positive for E gene were confirmed by performance of RT-qPCR for the detection of specific RdRP gene of SARS-CoV-2 using LightMix® Modular SARS-CoV-2 RdRP (TIB MOLBIOL) using similar PCR conditions as described above.

**Table 1:**
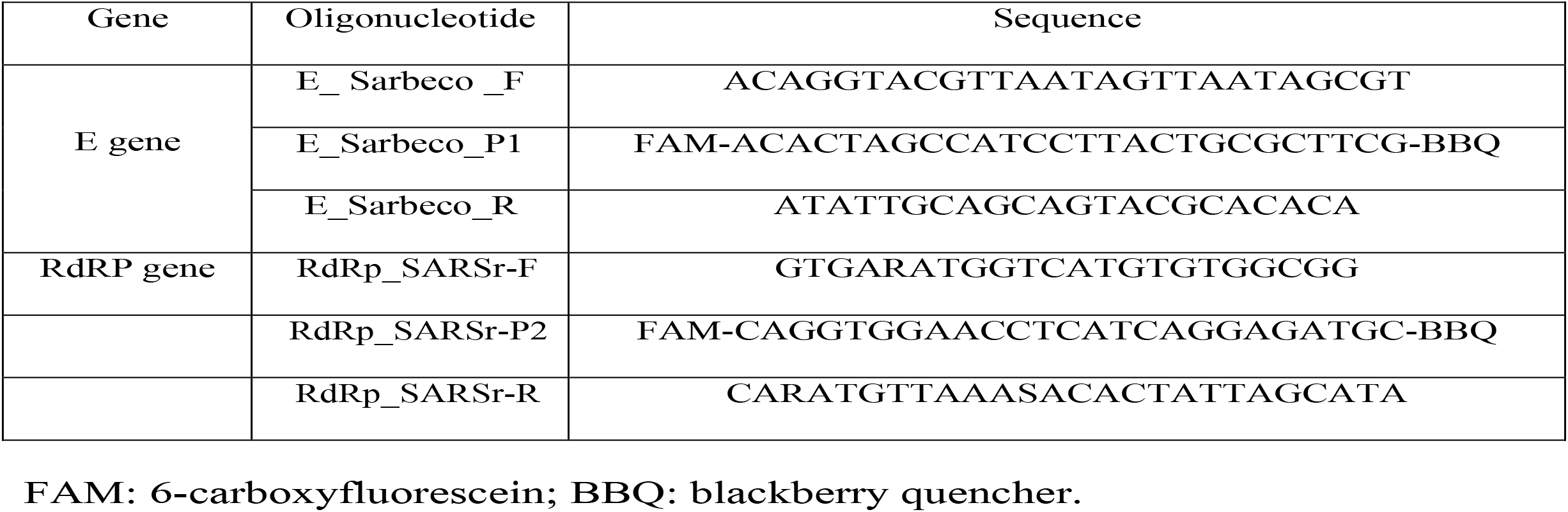
Primers and probes for the RT-qPCR.

### POOLING OF SAMPLES FOR THE RT-qPCR

RNA samples that were obtained after extraction were randomly pooled initially in pools of 2, 4, 6, 8, 16 RNA elutes on a 96 well plate. The best results matching with individual RNA test (ID) was seen in pools of 2, 4 and 8 samples. Subsequently, pools of 8 RNA elutes on a 96 well plate were used as shown in figure 1.Both ID and pooled RNA RT-qPCR for the screening E gene was done in the same plate (figure 1).5 µl of the RNA sample was taken for the PCR and pooling of 5 µl (8×5= 40 µl) was done for the pooled test. After thorough mixing, 5 ul of the pooled RNA was taken for the pooled PCR. The volume of RNA sample was kept similar in both the ID as well as pooled PCR so that assay sensitivity is not affected. ID PCR and pooled PCR was performed in the same run, keeping the entire conditions uniform.

**Figure 1:**
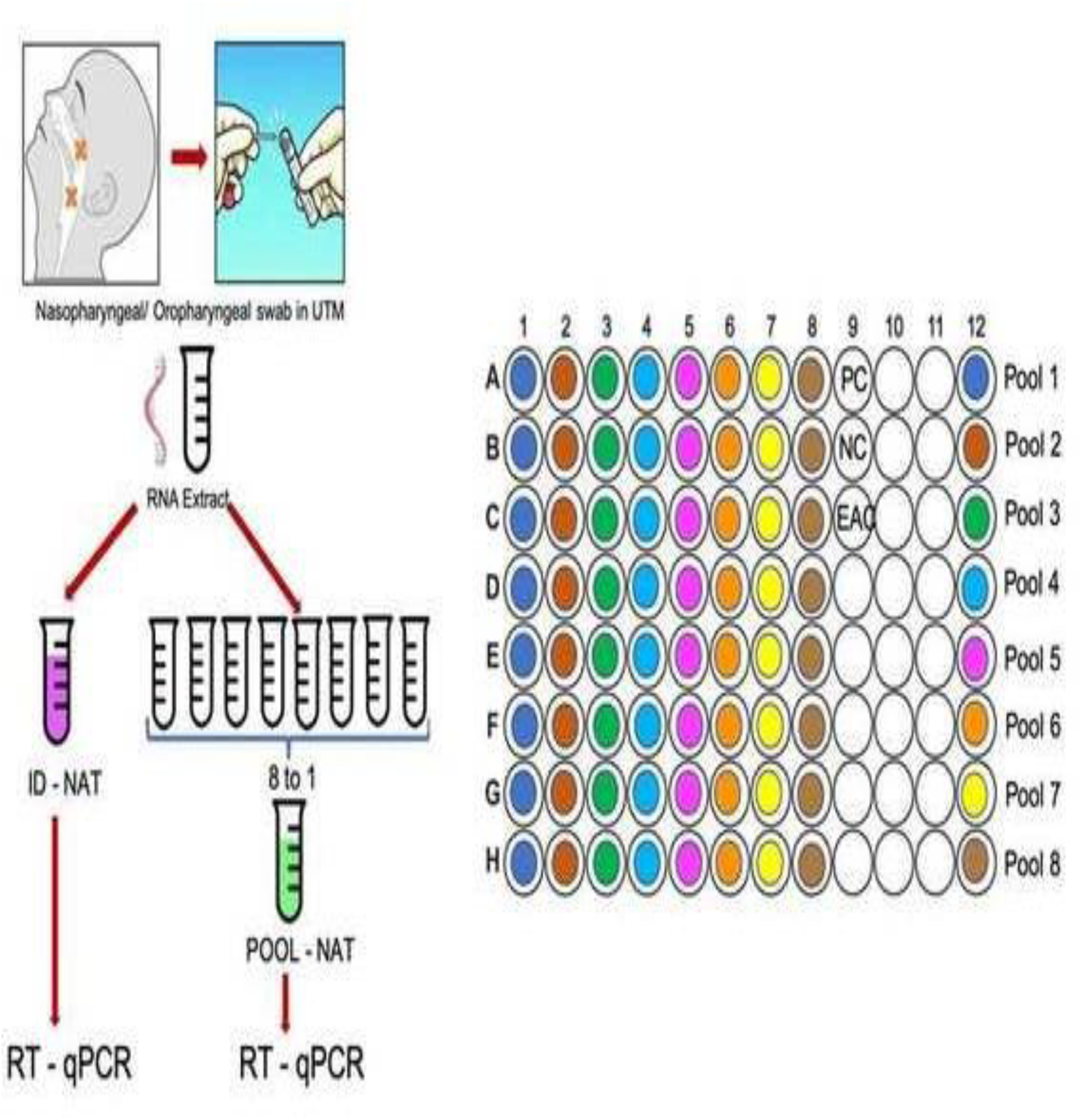
Scheme for pooling the samples on a 96 well plate.

Results were seen on the ABI software and each reaction was read for E gene after confirmation of the performance of EAC as well as positive control and negative control results. Ct value for each positive test was recorded and as per the WHO criteria, sample with Ct value ≤ 40 were considered as positive. All initial E gene positive were confirmed as positive if RdRP gene was also detected with Ct value ≤ 40.

### STATISTICAL ANALYSIS

The data was compared for the ability of detection of E gene by ID test and pooled test using one sample paired t-test (Table 1). Data of Ct value is shown as mean ± standard deviations. The agreement between Ct values obtained for positive sample in one positive sample pool with ID test was done applying intra class correlation ICC followed by Bland and Altman graph. A P value of 0.05 or less was considered statistically significant.

### ETHICAL APPROVAL

The study was approved by the Institutional ethics committee and patient consent form was waived off because of the use of deidentified discarded RNA samples.

## RESULTS

### POOL TEST Vs ID TEST RESULTS

Out of 280 samples that were tested, 40 were positive and 240 negative for SARS CoV-2 E gene with a positivity rate of 16.7% (95 CI; 12.2%-22.0%).All 40 were also positive for RdRP gene. Results were communicated to the patients as per the ID test results obtained.

All the clinical details were de-identified, delinked and kept anonymised as per the Government of India data safety policy. Overall 35 pools were made from these samples. On comparing the performance of pool test with ID test pools concordance in performance was seen in 34 pools. Overall sensitivity of the pool test keeping ID test as gold standard was 95.4%, specificity 100%, positive predictive value 100% and negative predictive value of 92.86%.

There were 13 pools where all the ID tests were negative and pool results were also negative. In 22 pools one or more positive sample was detected in the ID test (Table 2). Pools were further classified based on the number of positive samples present in them. There were 11 pools with 1 positive sample in each pool,5 with 2 positive sample in each pool,5 with 3 positive samples in each and 1 with 4 positive samples in it.21 out of 22 (95.4%) pools could correctly identify the positive test. In 1 pool which had 1 positive and 7 negative samples, the ID test result showed delayed Ct at 39, therefore it was missed. The sample was however positive for the RdRP gene and was given a positive result.

**Table 2:** Comparison between the mean Ct values of individual test and pool test in different pool combinations.

| Pool of 8 RNA samples |  | E gene detection |  |  |  |
| --- | --- | --- | --- | --- | --- |
| Pool combinations | Total Number of Pools | Not detected in number of pools | Mean of ID test Ct value (± SD) | Mean of Pool test Ct value (± SD) | P value |
| 1 positive + 7 negative | 11 | 1 | 33.1 (±3.3) | 35.6 (±3.3) | 0.041 |
| 2 positive + 6 negative | 5 | 0 | 30.4 (±3.7) | 33(±3.1) | 0.134 |
| 3 positive + 5 negative | 5 | 0 | 34.2 (±3.7) | 34.9(±2.6) | 0.581 |
| 4 positive + 4 negative | 1 | 0 | 31.7 (± 4.6) | 31.7 | - |
| 8 negative | 13 | 13 | NA |  | - |
| **Total** | **35** | **14** |  |  |  |

**Table 3:**
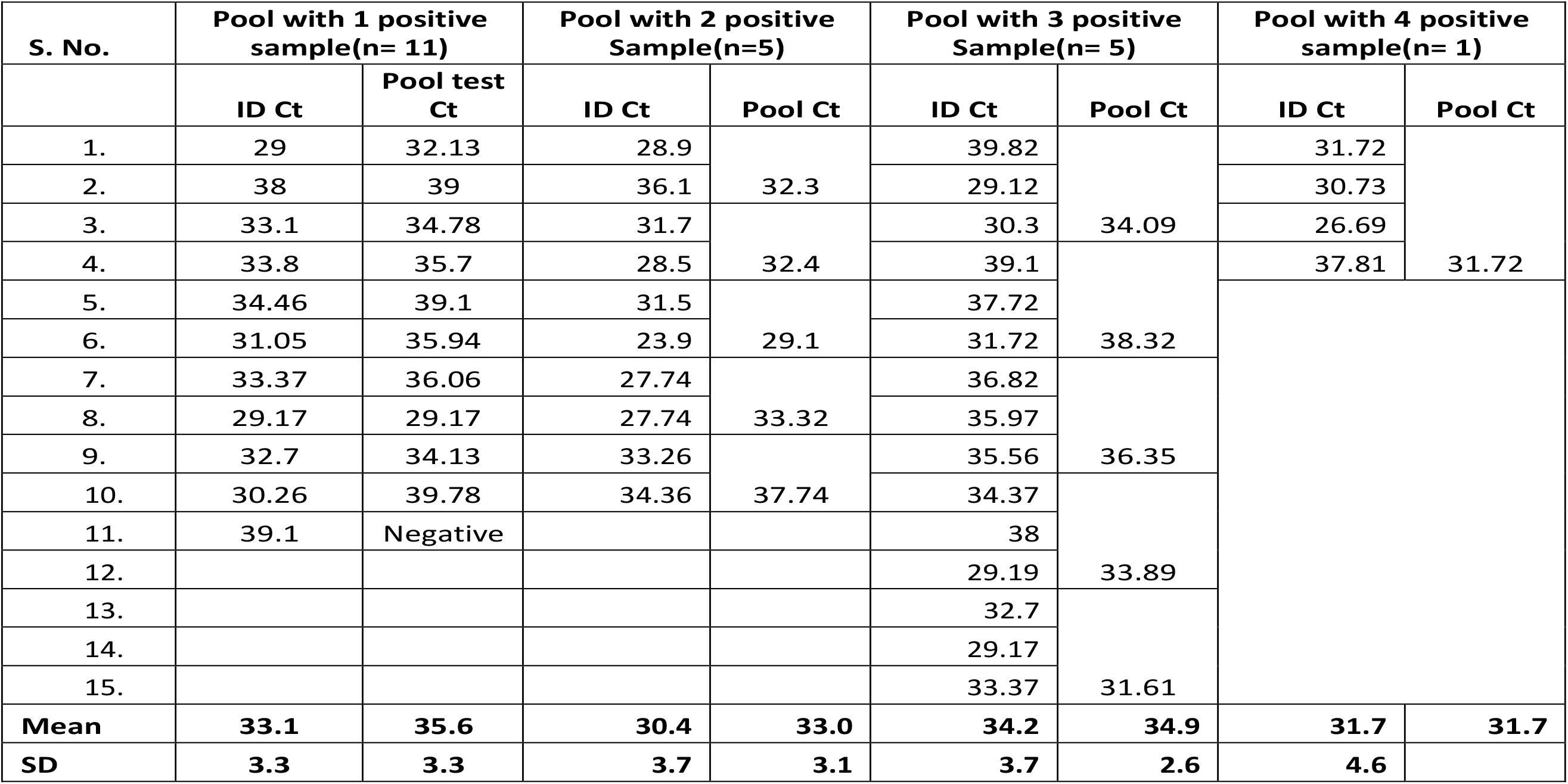
Comparison of individual test Ct value with pool test Ct value in the positive samples pool.

### EFFECT OF POOLING ON THE CT VALUE OF THE POSITIVE RESULT

In a positive test apart from getting a fluorescence curve the cycle number at which the fluorescence starts is also important, this is measured as Ct value. WHO has defined the criteria that any test which gives fluorescence after 40 cycles should not be considered as positive [9]. We wanted to see the effect of dilution on the Ct value of the positive result. Ct values of positive samples in ID test were compared with Ct values in pool test (Table 2).The pools having only 1 sample positive in them, the Ct values were similar to the ID test (p value =0.04).In pools with more than 1 positive sample the mean Ct value of positive as ID was compared with Mean Ct in pool, the overall mean CT value of ID was 32.68 while in a pooled testing it was 34.24 only an increase in Ct value of 1.56 very much with in the reporting criteria of being called as positive. This clearly shows that the dilution did not alter the Ct value of the positive result and a positive will be reported as positive despite pooling.

### EFFECT OF NUMBER OF POSITIVE SAMPLES IN A POOL

We tried to look at the effect of presence of number of positive samples in a pool. As the number of positive samples in a pool increased the difference in the mean Ct value decreased (figure 2). More the number of positives in a pool, more accurate the result of the pool testing. When there were 4 positive samples in a pool, the mean ID Ct value were identical to mean pool Ct (Table 2).

**Figure 2:**
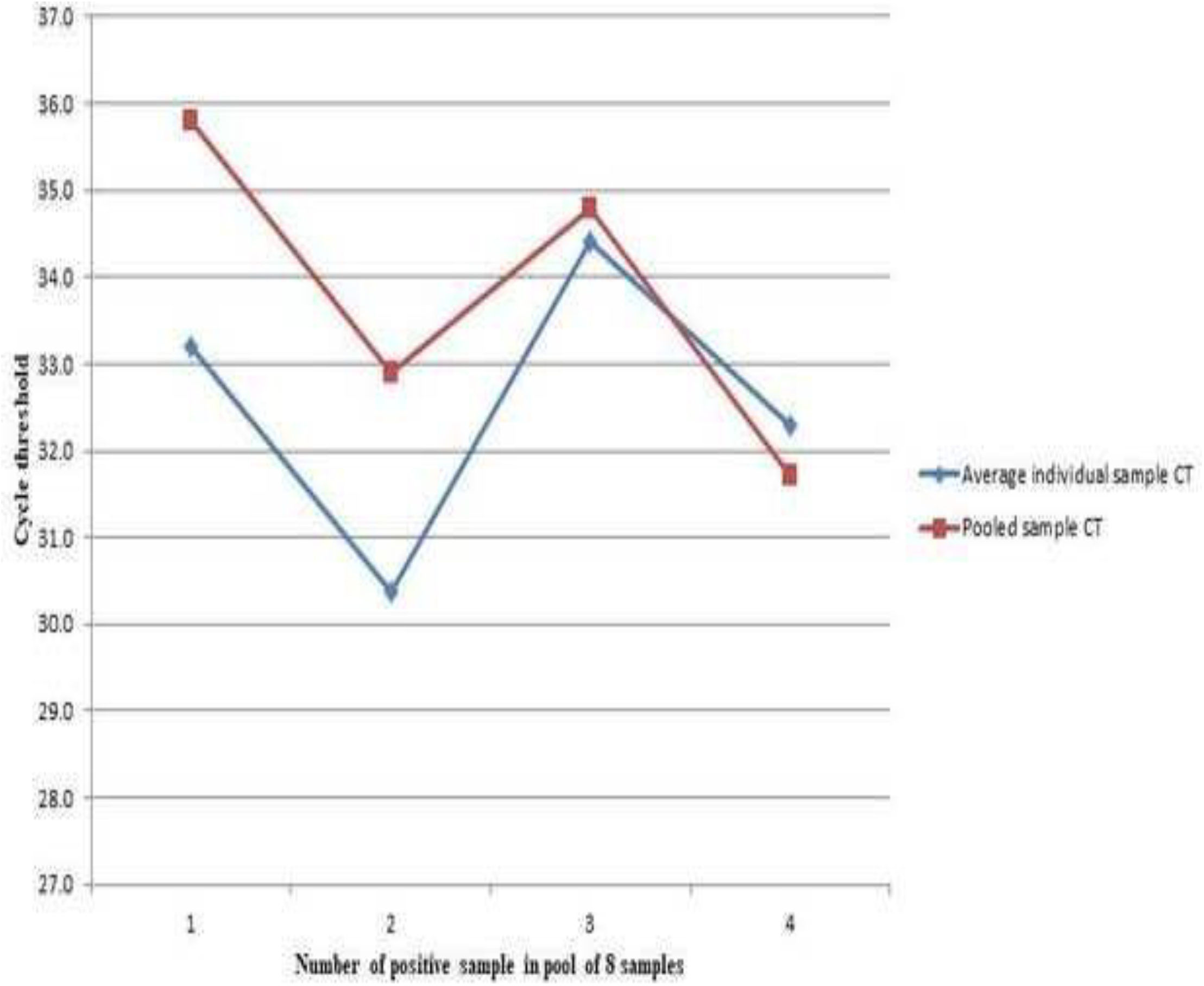
Effect of number of positive samples in a pool.

On evaluating the performance of pools with only 1 positive sample in it, a bland and altman graph (figure 3) showed an intra class agreement of 57.3% [(−29.4% to 88.7%), p=0.024]. The reliability coefficient was found to be 74.5%. All tests were within 2 SD limit except 1. The limit of biasness were -8.4 to 2.3.

**Figure 3:**
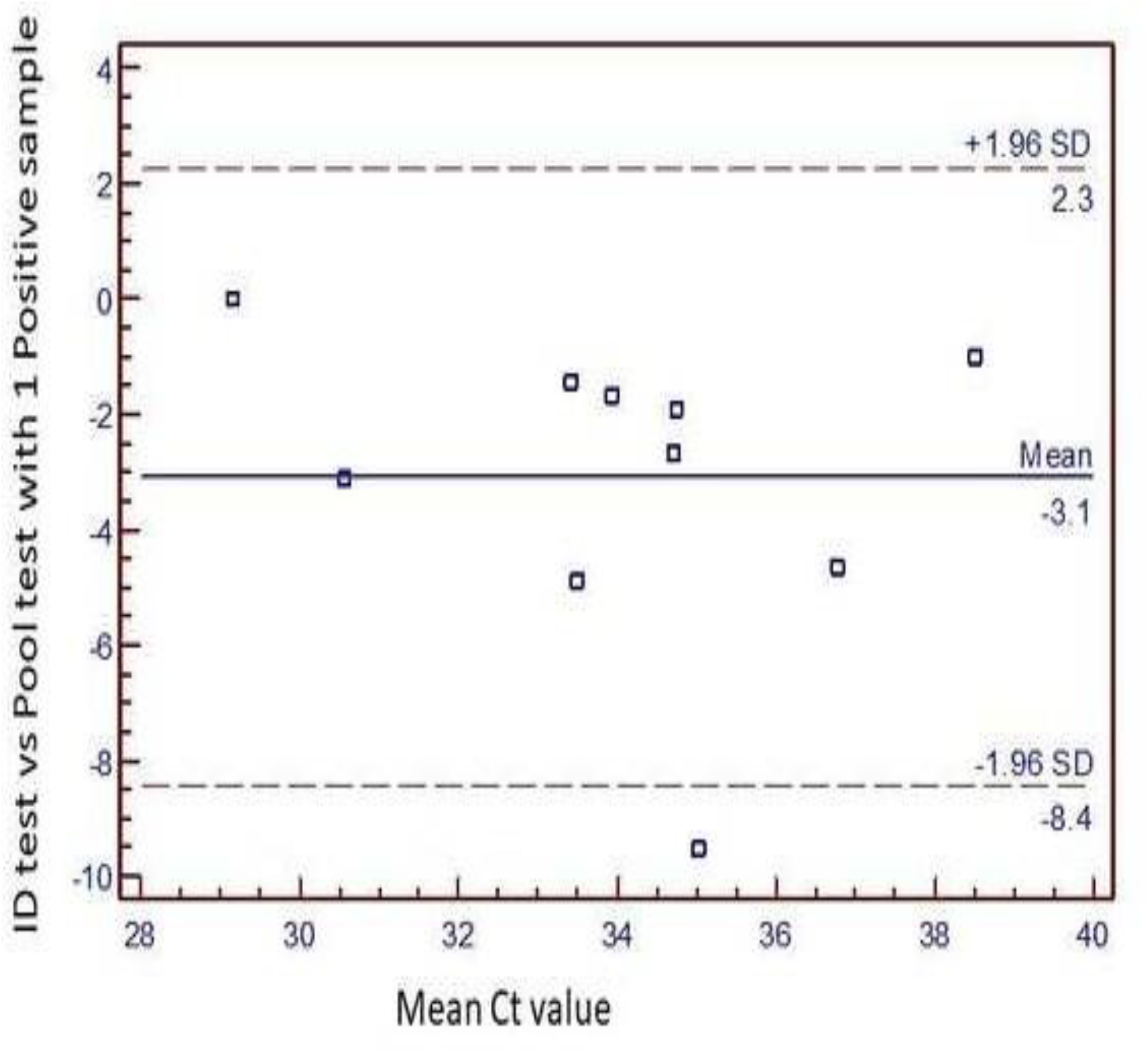
Performance of single positive sample pool.

## DISCUSSION

This novel study demonstrates that pooling of RNA samples can be done without loss of sensitivity and specificity. The results show that pooled test and ID test for the SARS-CoV-2 PCR, has demonstrated that pool testing with 8 RNA samples can easily detect even upto a single positive sample with Ct value as high as 38. The present study also showed that the results of pool testing is not affected by number of positive samples in a pool. Pool testing thus can be easily applied for faster PCR results. The data encourages wide spread of the technique for the diagnosis of SARS-CoV-2 infection as it saves time and expensive PCR reagents during the constrained shipping and supplies. Pooled testing has been used in many infectious diseases as a simple cost effective method to enhance the speed of diagnosis especially when large samples have to be tested during epidemic screening [10], pooling has been proven to work for RT-qPCR [7] [11], a time-consuming step for which the reagents are expected to be in short supply [12].

The WHO declared COVID-19 as a global pandemic on March 11^th^, 2020 [13]. As aptly described by the WHO director general, the *mantra* of “detect, protect and treat” should be followed to break the chain of transmission of SARS-CoV-2 [14]. Early diagnosis and prompt treatment can substantially reduce the number of prospective cases. Hence, laboratory diagnosis of SARS-CoV-2 holds the key in containing and restricting the COVID-19 pandemic. PCR remains the gold standard for the diagnosis of COVID-19 infection especially in symptomatic individual, but limited supply of diagnostic kits and reagents are major bottle neck in expediting testing of COVID-19 in the community [4]. Furthermore, mass testing of samples should be done to escalate testing and even extend to asymptomatic cases [15]. India a country of over 1.3 billion population [16] testing for COVID-19 has been a great challenge. Government of India through the Indian Council of Medical Research (ICMR) has established a robust network of laboratories in the form of Viral research and diagnostic laboratories (VRDL) [17]. These VRDLs have taken a lead in the real time diagnosis of COVID-19 in India. Still to match up to such a humongous population of India, we need some fast and rapid solutions to intensify the testing. Hence a testing method where a large number of samples can be tested and consuming minimal testing kits and reagent is the need of the hour. Pooled testing can solve this problem. In another study on pool testing of samples maximum number of pooling of samples was assessed and in this study they found that pooling of samples upto a pool of 64 samples together can be done without effecting the results [18].

In the present study, the pool size was limited to 8 based on the preliminary analysis done by us on determination of pool size (unpublished data). Pooling of samples can be done at various levels, prior to RNA extraction that is at the time of sample collection, putting the nasopharyngeal/ oropharyngeal swabs into a common viral transport media(VTM). By doing so the major bottle neck of RNA extraction can be removed. Pooling can also be done by pooling the VTMs and doing common extraction for pooled VTM samples, here the limitation is that in case a pool is positive individual sample collection has to be done at the patient level. Pooling RNA samples are easy to do in a laboratory as any pool turns out to be positive ID test can be performed and result can be declared.

It can be argued that the sample size of our study is small. However, in the interest of widespread need to enhance testing facilities and capabilities, the present study makes a seminal contribution. In future, mathematical and computational model can be applied to design appropriate pool size and increase the pool size further for SARS-CoV-2 testing. Pooling of samples is essentially important in monitoring the infection in cohesive groups such as quarantine facilities, health care workers, community surveillance and diagnosing the asymptomatic cases. The infection load in these groups may be low but even a single positive case amongst such groups can activate the surveillance system and quarantine the affected group and prevent the further spread in the community.

## CONCLUSION

As the COVID-19 pandemic spans across the globe, implementation of expanded testing in larger population groups is the only way out. We recommend that testing the pooled samples, if done properly, is reliable, and can take the fight of detecting COVID-19 to the next level at the earliest.

## Data Availability

Lab based

## CONTRIBUTORS

EG, SKS conceived and designed the study. AP, AK, RA, KR, VM, MK, SD did the literature search. Laboratory data were collected by AK. Data analysis was done by GK. EG interpreted the data. EG, AP and SKS drafted and prepared the manuscript.

## Acknowledgements

We acknowledge all the technical staff involved in testing and ICMR for providing the Primers and probes for the conduct of RT-q PCR.

